# Mendelian randomization analysis identifies a causal effect of Streptococcus salivarius on DR mediating via the level of host fasting glucose

**DOI:** 10.1101/2023.12.19.23300249

**Authors:** Jingjing Li, Gongwei Zheng, Dingping Jiang, Chunyu Deng, Yaru Zhang, Yunlong Ma, Jianzhong Su

## Abstract

**Background:** Diabetic retinopathy (DR) is one of leading causes of vision loss in adults with increasing prevalence worldwide. Increasing evidence has emphasized the importance of gut microbiome in the etiology and development of DR. However, the causal relationship between gut microbes and DR remains largely unknown.

**Methods:** To investigate the causal associations of DR with gut microbes and DR risk factors, we employed two-sample Mendelian Randomization (MR) analyses to estimate the causal effects of 207 gut microbes on DR outcomes. Inputs for MR included Genome-wide Association Study (GWAS) summary statistics of 207 taxa of gut microbes (the Dutch Microbiome Project) and 21 risk factors for DR. The GWAS summary statistics data of DR was from the FinnGen Research Project. Data analysis was performed in May 2023.

**Results:** We identified eight bacterial taxa that exhibited significant causal associations with DR (FDR < 0.05). Among them, genus *Collinsella* and species *Collinsella aerofaciens* were associated with increased risk of DR, while the species *Bacteroides faecis*, *Burkholderiales bacterium_1_1_47*, *Ruminococcus torques, Streptococcus salivarius*, genus *Burkholderiales_noname*, and family *Burkholderiales_noname* showed protective effects against DR. Notably, we found that the causal effect of species *Streptococcus salivarius* on DR was mediated through the level of host fasting glucose, a well-established risk factor for DR.

**Conclusions:** Our results reveal that specific gut microbes may be causally linked to DR via mediating host metabolic risk factors, highlighting potential novel therapeutic or preventive targets for DR.

## Introduction

Diabetic retinopathy (DR) is a vision-threatening complication in patients with diabetes (both type 1 and type 2) and has been one of the leading causes of vision loss in adults aged 20–74 years [1]. Among individuals with diabetes, the overall prevalence of DR is estimated to be 34.6% [1]. According to the Global Burden of Disease Study [2], while the prevalence of blindness for other causes consistently showed a regional decrease between 1990 and 2020, the prevalence of DR increased in many regions, especially Asia, sub-Saharan Africa, and high-income North America. Given that people with diabetes live increasingly longer, the number of people with diabetic retinopathy and resulting vision impairment is expected to rapidly rise, reaching 160.50 million by 2045 from 103.12 million in 2012 [3].

The well-established major risk factors for DR contain hyperglycemia, hypertension, and dyslipidemia. A pooled analysis using 22,896 participants from 35 studies in the U.S., Australia, Europe, and Asia has summarized that the prevalence of DR increased with diabetes duration, HbA1c, and blood pressure [1]. The Singapore Malay Eye Study showed that independent risk factors for DR were longer diabetes duration, higher hemoglobin A1c, hypertension, and higher pulse pressure [4]. In the Chinese type 2 diabetic patients’ study, hyperlipidemia, higher VLDL, and higher triglyceride were independently associated with the increased risk of DR [5]. In addition, inflammatory markers and cytokines, such as CRP, IL6, IL8, IL16, and TNF-α, were also implicated to be associated with the development of DR [6–9].

Extensive studies have highlighted the importance of oxidative stress induced by hyperglycemia on the development of DR [10]. Abnormal cellular signaling between the neuronal and vascular retina, increased retinal vascular permeability, neovascularization and altered neuronal functions were involved in the etiology of DR. A variety of molecules and neurovascular signaling pathways have been reported to be associated with DR risk, such as vascular endothelial growth factor A (VEGFA), kinin–kallikrein system, angiopoietin-like 4 (ANGPTL4) and Leucine-rich α2-glycoprotein 1 (LRG1) [11].

Recently, gut microbiome dysbiosis is proposed to be associated with eye diseases, including uveitis, glaucoma, age-related macular degeneration, and DR, deriving the concept of gut-eye axis [12]. Multiple lines of evidence have shown that there exist a remarkable involvement of aberrant gut microbiota in ocular diseases based on on human and mouse models [12]. For instance, translocation of peptidoglycan derived from cell wall of gut microbes to retina was identified to activate TLR2-mediated MyD88/ARNO/ARF6 signaling pathway and promote DR [13]. Moreover, reconstruction of gut microbiome by antibiotic therapies or intermittent fasting have been distinguished to prevent the development of DR [14]. Despite evidence that gut microbiome affects DR, there is paucity of knowledge regarding which specific species or clades of gut microbes are causal for DR, and how these causal gut microbes influence DR.

Genome-wide association study (GWAS) of microbe abundances have identified genetic variants that are associated with microbes [15], providing a promising opportunity to examine the causal effects of microbes on DR using Mendelian Randomisation (MR) [16]. In this study, we investigated the causal associations of DR with gut microbes and DR risk factors using two-sample Mendelian Randomization framework. We collected and curated largest and most comprehensive GWAS summary statistics of 207 taxa of gut microbes and 21 risk factors for DR. Causal mechanisms were explored using mediation analyses, revealing the role of blood biochemical indicators (e.g., blood glucose) in mediating the effects of gut microbes on DR. In summary, our systematic MR analysis provides valuable novel insights into the complex interplay among the gut microbiome, metabolic risk factors, and DR.

## Methods

### Data source

Microbe GWAS summary data based on the largest genome-wide association study (GWAS) of 207 gut microbial taxa (Supplementary Table S1) were downloaded from the Dutch Microbiome Project [15]. The Dutch Microbiome Project has launched and reported the large-scale GWAS of gut microbiome composition in 7,738 volunteers from Netherlands by applying metagenomic sequencing, which allows for bacterial identification at species-level resolution.

For DR, we utilized summary statistics from the FinnGen Research Project, which is the leading biobank-based genomic research project with more than 1,900 diseases aiming to include 500,000 Finland participants (n = 218,792 participants in data release 5) [17]. Broad diabetic retinopathy phenotypes were extracted from phenotype documentation of FinnGen database release 5 (https://www.finngen.fi/en/researchers/clinical-endpoints). After stringent quality control and phenotype screening, we included 3 DR-relevant phenotypes, whose phenotype codes were H7_RETINOPATHYDIAB (denoted as DR1), DM_RETINOPATHY_EXMORE (denoted as DR2) and DM_RETINOPATHY (denoted as DR3). DR1 was from the category of “VII Diseases of the eye and adnexa”, while DR2 and DR3 were from the category of “diabetes endpoints”. Overall, 3,646 cases and 203,018 controls were included in DR1, 14,584 cases and 176,010 controls were included in DR2, 14,584 and 202,082 controls were included in DR3 (Supplementary Table S2).

### Metabolic Risk factors for DR

The development of DR is related to many metabolic risk factors and dysfunctions [18]. Hyperglycemia, hypertension, and dyslipidemia were reported to be major risk factors for DR [1, 4]. Thus, we included GWAS of plasma metabolic variables representing the three major risk factors from the FinnGen project, which were fasting insulin (FI), fasting glucose (FG), HbA1c, 2h glucose after an oral glucose challenge (2hGlu), triglyceride (TG), very low-density lipoprotein (VLDL), high-density lipoprotein (HDL), low-density lipoprotein (LDL), hypercholesterolaemia, high cholesterol, systolic blood pressure (SBP), and diastolic blood pressure (DBP). In addition, body mass index (BMI) [19] and other previous reported DR risk factors, including C-reactive protein (CRP) [6], IL6 [7], CCL5 [20], IL8, IL16 [8], and nerve growth factor (NGF) [21], were also included in this study. For these included putative DR-relevant risk factors, GWAS summary statistics were restricted to European populations.

### Mendelian Randomization strategy

The principle of MR is using genetic variation as instrumental variable to estimate the causal relationship between exposure and outcome [22]. The MR approach relies on three key assumptions: (i) the genetic variants used as instrumental variables (IVs) are associated with the preset exposure variable. (ii) there are no unmeasured confounders affecting the associations between genetic variants and outcomes; and (iii) the genetic variants affect the outcome only through changes in the exposure, without exhibiting pleiotropy.

In this study, two-sample MR method [22, 23] was applied to evaluate causal relationship between predefined exposure and outcome variables. SNPs reaching significance threshold (p < 1e-05) were extracted from GWAS summary statistics of exposure and used as instrumental variables (IVs). To ensure statistical independence across SNPs, we conducted LD clumping with a cut-off of r^2^ < 0.1 within a 1Mb window. Statistics of these IVs were subsequently extracted from GWAS summary statistics on outcome. After harmonising the direction of estimates of SNP-exposure and SNP-outcome associations, MR estimates were generated to assess the effect of genetic liability of exposure to outcome. To obtain the MR causal effect estimates, Wald’s ratio method was applied when there is only one (IVs) available for exposure. The inverse variance weighting (IVW) method in fixed-effect framework was applied if the number of IVs is between 1 and 3. For the case that the number of IVs surpass 3, we utilized the inverse variance weighting (IVW) method in multiplicative random-effects framework [24]. The IVW method combining with multiplicative random-effects model was able to handle the dispersion of effect estimates due to pleiotropy. To avoid violating the MR assumption, we performed three sensitivity analyses to assess the robustness of the results. Specifically, heterogeneity was estimated by the Cochran Q test [25]. Horizontal pleiotropy was estimated using MR-Egger’s intercept [26], and influential outlier IVs due to pleiotropy were assessed using MR Pleiotropy Residual Sum and Outlier (MR-PRESSO) [27].

### Systematic MR screening for putative causal gut microbes and risk factors of DR

At the first stage, we applied the two-sample MR strategy to evaluate causal relationship between gut microbes and DR by defining gut microbes as exposures and DR as outcome (Gut microbes → DR). We employed the two-sample MR strategy for all gut microbial taxon - DR pairs. All statistical tests were two-sided and adjusted for multiple hypothesis testing (Benjamini-Hochberg method). For each bacterial GWAS summary dataset, we calculated the MR causal effect estimates by combining it with each of the three DR GWAS summary statistics. We considered the bacteria to be causal for DR if (i) at least one of the three causal associations reach significant threshold (FDR < 0.05); and (ii) direction of all three MR estimates are identical. Bacteria taxon that passed the screening criteria were included in the following MR analyses. Next, we applied the two-sample MR analyse on putative DR-relevant risk factors by defining them as exposures and DR as outcome (DR risk factors → DR). Finally, the MR strategy was implemented to infer causal relationships between putative causal gut microbes identified in the first stage and DR risk factors derived form the second step (Gut microbes → DR risk factors). The MR strategy and screening criteria for each step were consistent with the first stage. The MR analyses, Cochran Q, and MR-Egger sensitivity analyses were conducted using R package TwoSampleMR (Version: 0.5.6). MR-PRESSO test was performed using R package MRPRESSO (Version: 1.0). GWAS data was processed by R packages ieugwasr (Version 0.1.5), reshape2 (Version 1.4.4), and dplyr (Version 1.0.7). Plots were visualized using R packages ggplot2 (Version 3.4.0), circlize (Version 0.4.15), and ComplexHeatmap (Version 2.13.1). All statistical analyses were performed in R 4.1.4 (www.R-project.org).

### Mediation analysis

For “gut microbe – DR risk factor – DR” triplets that passed the MR screening, we performed mediation analyses to quantify the causal effects of gut microbes on DR via the risk factors. The total effect of gut microbes on DR was assessed by the primary MR (Gut microbes → DR). The indirect effect was estimated by two-step MR. In first Step, univariable MR model was employed to estimate the effects of the gut microbes on the DR risk factors (Gut microbes → DR risk factors). It involved assessing the causal relationship between the gut microbes and individual risk factors for DR. In second step, multivariable Mendelian randomization (MVMR) was carried out to estimate the effect of DR risk factor on DR (DR risk factors → DR), adjusting for the effect of corresponding gut microbe. It allowed us to evaluate the specific impact of the risk factors on DR, taking into account the potential confounding influence of the gut microbe. We used the product of these two estimates to calculate the indirect effect, as analogue to a previous study [28]. Standard errors and confidence interval (CI) of the indirect effect were estimated by using the Delta method [16, 29]. The proportion mediated effect was yielded from dividing the indirect effect by the total effect.

## Results

### Causal gut microbes for DR

Figure 1 shows the workflow of the current study. A total of 207 gut microbes were tested for pinpointing causal relationships with DR (Supplementary Table S1 and S3). According to our screening criterion (see Methods), we identified 8 bacterial taxa that were causally associated with DR (Fig. 2, Supplementary Fig. S1 and Supplementary Table S4), namely, *g_Collinsella, s_Collinsella_aerofaciens, s_Bacteroides_faecis, s_Burkholderiales_bacterium_1_1_47, f_Burkholderiales_noname, g_Burkholderiales_noname, s_Ruminococcus_torques,* and *s_Streptococcus_salivarius*. Among these taxa, *g_Collinsella* (OR per 1-SD higher bacterial abundance [95% CI] = 1.25 [0.03, 0.42] for DR2; 1.20 [0.09, 0.28] for DR3) and *s_Collinsella_aerofaciens* (OR [95% CI] = 1.24 [0.11, 0.33] for DR2; 1.09 [0.002, 0.16] for DR3) were associated with a higher risk of DR, while other taxa were associated with a lower risk of DR. This is consistent with previous opinion that *Collinsella* exerted adverse effects on human health [38] and contributed to the development of insulin resistance and diabetes [39].

**Figure 1.**
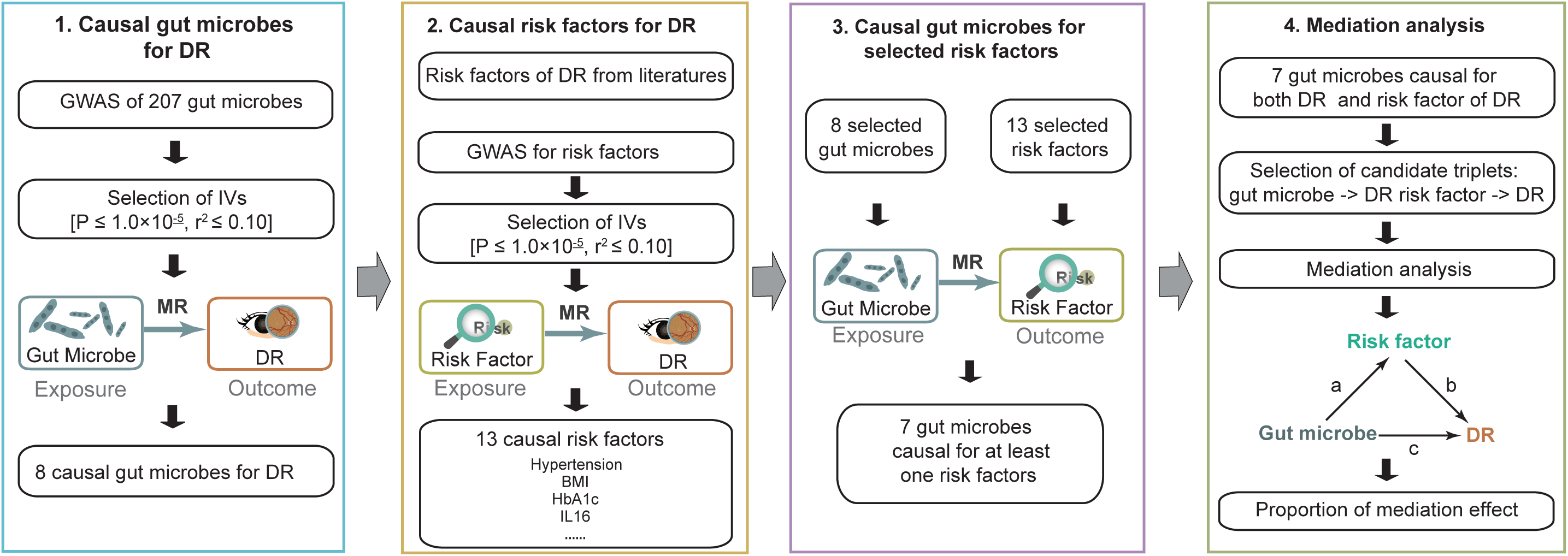
Overview of this MR study. Four O-link panels were applied to measure associations among gut microbes, DR risk factors and DR. At the primary MR step, genetic variants associated with gut microbes were identified based on results from their corresponding GWAS. Genetic variants that passed significant threshold were then used as instrumental variants to test their relationship with DR. In the second step, genetic variants associated with DR risk factors was extracted from corresponding GWAS to test MR causal relationship between DR risk factor and DR. Then, MR was applied to measure causal relationship between gut microbes that passed significant threshold in the primary MR and DR risk factors screened from the second step. Finally, mediation analyses by two-sample MR were performed for the gut microbes that were causally associated with DR risk factors and DR.

**Figure 2.**
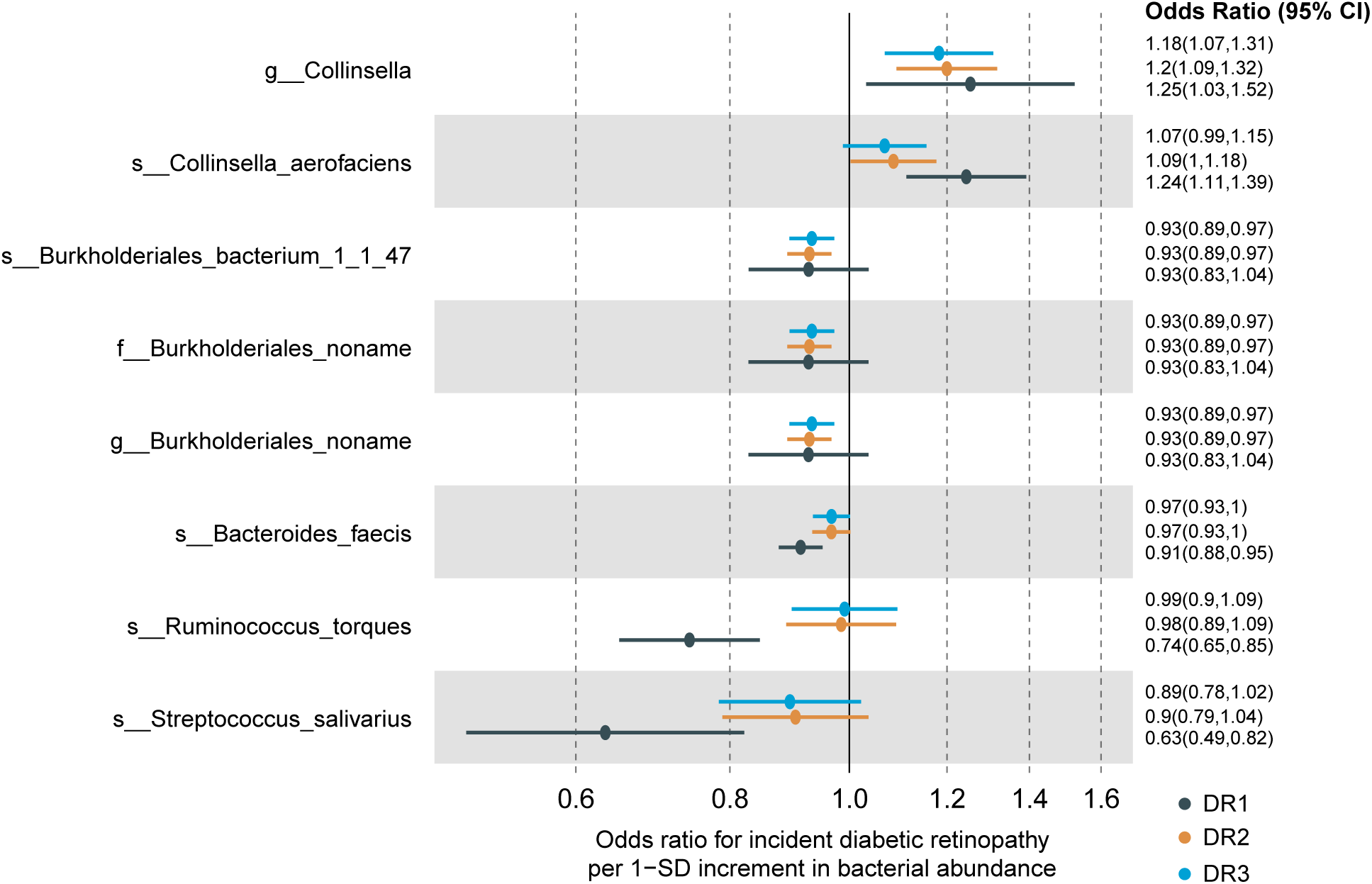
Effects of eight potential causal gut microbes on DR. MR analyses of the effect of gut microbes on DR. The dots are the causal estimates on the OR scale, and the whiskers represent 95% confidence intervals for the ORs. P values were determined from the inverse-variance-weighted two-sample MR method.

Results of sensitivity analyses confirmed the robustness of the MR analyses for these taxa (Supplementary Table S4). As tested by Cochran Q statistics, there was no evidence for heterogeneity (P_Q-stat_ > 0.05) or horizontal pleiotropy (MR-Egger test, P_Egger-Intercept_ > 0.05; MR-PRESSO global pleiotropy test, P_GlobalTest_ > 0.05, Supplementary Table S4).

### Causal risk factors for DR

To uncover potential DR risk factors that mediate the causal effects of gut microbes on DR, we first applied the two-sample MR strategy to screen risk factors that were causally associated with DR. For each of these risk factors included in this study, IVs were extracted from corresponding GWAS summary statistics restricted to European populations (Supplementary Table S5). There were 12 risk factors identified to be significantly causally associated with DR-relevant phenotypes. Among them, FI, Hypertension, FG, BMI, HbA1c, 2hGlu, TG, SBP, and DBP exhibited prominent associations with increased risk of DR, while IL16, LDL, and HDL were significantly associated with lower risk of DR (Fig. 3, Supplementary Fig. S2, and Supplementary Table S6). FI showed the strongest effect on DR (OR [95% CI] = 2.51 [1.54, 4.10] and 1.78 [1.38, 2.30] for DR1 and DR2, respectively), followed by hypertension (OR [95% CI] = 2.14 [1.73, 2.65] and 2.13 [1.38, 3.31] for DR1 and DR2, respectively).

**Figure 3.**
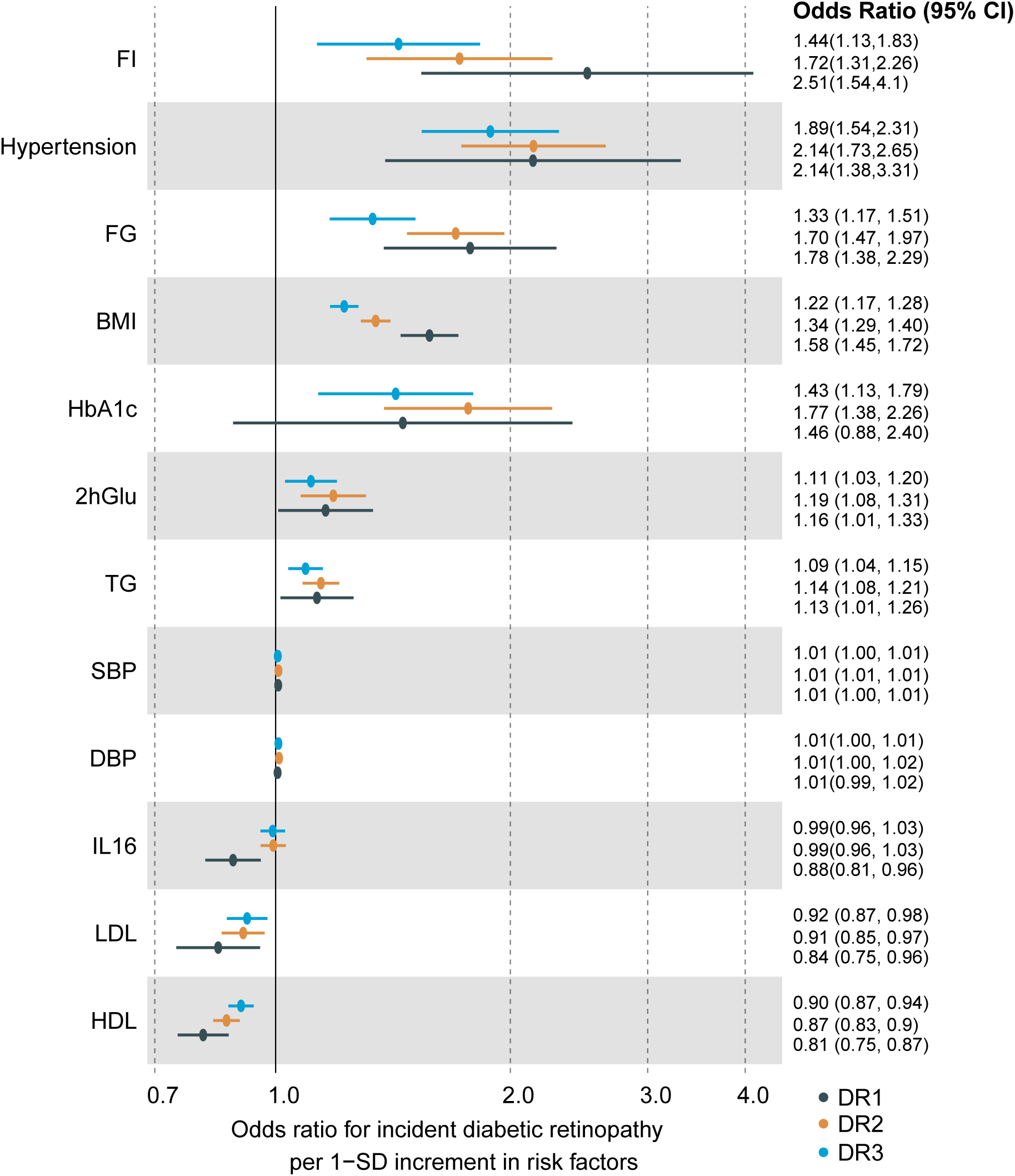
Causal effects of risk factors on DR. MR analyses of the effect of risk factors on DR. The dots are the causal estimates on the OR scale, and the whiskers represent 95% confidence intervals for the ORs. P values were determined from the inverse-variance-weighted two-sample MR method. FI: fasting insulin, FG: fasting glucose, 2hGlu: 2h glucose after an oral glucose challenge, TG: triglyceride, HDL: high-density lipoprotein, LDL: low-density lipoprotein, SBP: systolic blood pressure, DBP: diastolic blood pressure, BMI: body mass index.

Due to there exist significant heterogeneity (MR-PRESSO Global Test p < 0.05 or an MR-Egger Intercept p < 0.05) ( Supplementary Table S7), we leveraged the inverse variance weighting (IVW) method in multiplicative random-effects framework, which is preferred when there exist heterogeneity [30].

### Risk factors associated with gut microbes

To prioritize critical risk factors exhibiting causal associations with both DR and gut microbes, we subsequently performed two-sample MR of these 12 significant DR-relevant risk factors with 8 DR-related bacterial taxa. A total of eight significant pairs of gut microbe and risk factor were identified (FDR < 0.05), and five pairs showed suggestive associations (P < 0.05, Fig. 4 and Supplementary Table S8). Among these significant pairs, we found that Taxon *s_Streptococcus_salivarius* was associated with lower risk of 2hGlu (OR [95% CI] = 0.88 [0.82, 0.95]) and FG (OR [95% CI] = 0.96 [0.95, 9,98]). Taxa *f_Burkholderiales_noname*, *g_Burkholderiales_noname,*and *s_Burkholderiales_bacterium_1_1_47* were associated with lower risk of HbA1c (OR [95% CI] = 0.98 [0.979, 0.996]) and higher risk of IL16 (OR [95% CI] = 1.05 [1.01, 1.09]). The other five suggestive associations included lower SBP risk of *g_Collinsella* (OR [95% CI] = 0.742 [0.597, 0.924]) and *s_Collinsella_aerofaciens* (OR [95% CI] = 0.743 [0.594, 0.930]), lower DBP risk of *s_Collinsella_aerofaciens* (OR [95% CI] = 0.654 [0.457, 0.935]), higher IL16 risk of *s_Collinsella_aerofaciens* (OR [95% CI] = 1.107 [1.025, 1.196]) and lower LDL risk of *s_Ruminococcus_torques* (OR [95% CI] = 0.981 [0.966, 0.997]).

**Figure 4.**
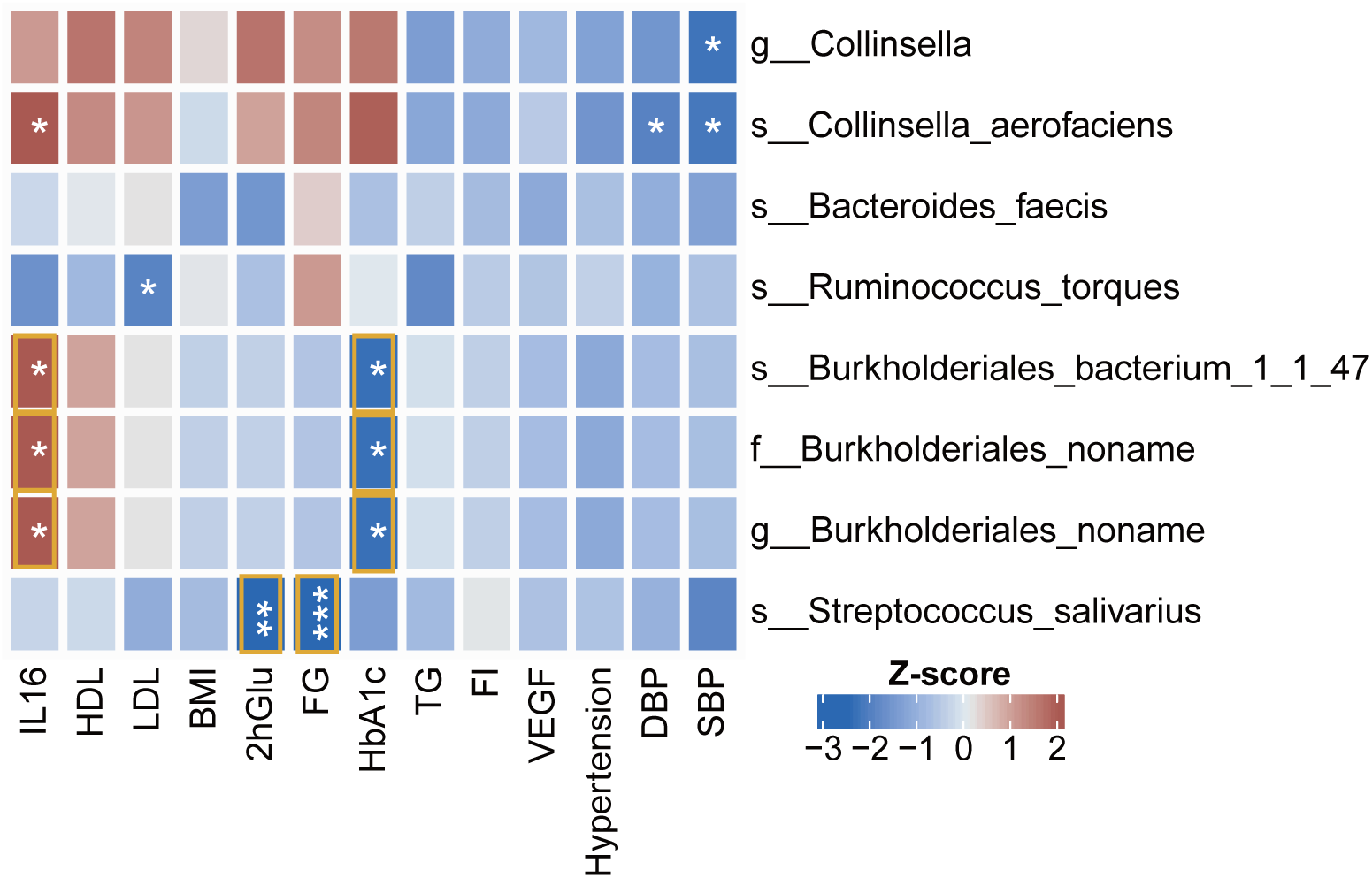
Effect sizes (Z-score) of eight potential causal gut microbes on causal risk factors for DR. MR analyses of the effect of gut microbes on DR risk factors. Colours in the heatmap represent the effect size (Z-score), with genetically predicted increased bacteria level associated with a higher risk of outcomes colored in red and lower risk of outcomes colored in blue. The darker the color the larger the effect size. * Indicates that the causal association passed the threshold of raw p value < 0.05. ** raw p value < 0.01. *** raw p value < 0.001. Orange box indicates that the causal association passed the threshold of FDR < 0.05.

### Mediation effect of gut microbes on DR via risk factors

Based on abovementioned two-sample MR strategy, eight “gut microbe-risk factor-DR” triplets were identified to show pairwise causal associations. Thus, we used these eight triplets to estimate the statistical significance of mediation effects and the proportion of the overall effects of gut microbes on DR that was mediated by corresponding risk factors (Supplementary Table S9). Through Delta method [16, 29], we obtained four triplets having mediation effects that passed the threshold of raw p-value < 0.05 (Fig. 5a). After multiple testing correction, one triplet, “*s_Streptococcus-salivarius* -> FG -> DR1”, reached significant threshold (FDR < 0.05, Fig. 5b). The proportion of mediation effect of *s_Streptococcus-salivarius* on DR via FG was 5.05%. For other three triplets with suggestive mediation effects (p-value < 0.05), we found that the causal effects of *s_Burkholderiales-bacterium-1-1-47/g_Burkholderiales-noname*/f_Burkholderiales-noname on DR were commonly mediated by HbA1c (Supplementary Table S9).

**Figure 5.**
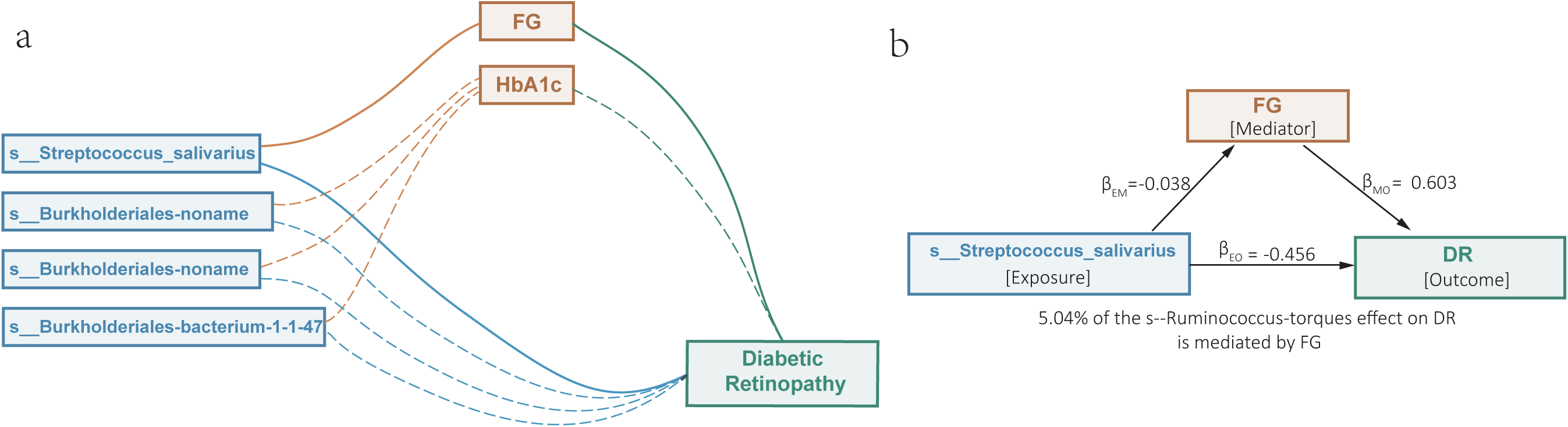
Mediation effects of gut microbes on DR via risk factors. Mediation analyses to quantify the effects of gut microbes on DR outcomes via risk factors. a Four “gut microbe – risk factor – DR” triplets that passed significant threshold (p value < 0.05) by mediation analyses. Solid lines indicate that mediation effect passed significant threshold of FDR < 0.05. Dotted lines indicate that mediation effect passed significant threshold of raw p value < 0.05. b Species Ruminococcus torques effect on DR mediated by fasting glucose. βEM effects of exposure on mediator, βMO effects of mediator on outcome, βEO effects of exposure on outcome.

## Discussion

In this study, to explore causal gut microbes for DR, we applied two-sample MR analyses to examine causal associations between gut microbes, risk factors, and DR. Our results highlighted that eight bacterial taxon and 13 metabolic factors were casually associated with DR. Further, mediation analysis supported that the causal effects of gut microbes on DR were partially mediated by metabolic factors that related to DR.

Accumulating evidence supports the connection between gut microbiome and retina diseases, which is also called “gut-retina axis” [12, 31]. Dysbiosis in gut microbiota was convinced to contribute to the development of diabetes mellitus (DM) and its microvascular complications, including DR. For T1DM, gut microbes were involved in the autoimmune mechanisms, such as regulating immune cell functions and production of anti-inflammatory cytokines [32]. In T2DM, the dysbiosis of gut microbiome promotes intestinal permeability, LPS translocation, hyperactivation of inflammatory responses, and dysregulation of insulin-related pathways, exacerbating the progression of insulin resistance [33].

Regarding DR, gut microbes could influence the homeostasis of retinal tissue through multiple metabolities. Oresic et al. [34] have reported that gut microbiota affects lens and retinal lipid composition by comparing lipidomic profiling of lens and retina of germ-free and conventionally raised mice. Trimethylamine N-oxide (TMAO) derived from gut microbes could be an important regulator of DR-related dyslipidemia [35]. Floyd and Grant [12] have summarized supporting evidence for gut-eye axis derived from murine models, highlighting the role of secondary bile acid produced by gut microbes, namely tauroursodeoxycholate (TUDCA), in preventing exacerbation of DR. In addition, peptidoglycan (PGN) synthesized by gut microbiota could translocate into the circulation and travel to the retina and exacerbate DR by modulating retinal endothelial cells [13]. Although previous studies have demonstrated the association of gut microbes with DR, evidence for causal relationship between them is still limited.

In this MR study, we found that genus *Collinsella* and species *Collinsella aerofaciens* were detrimental, while species *Streptococcus salivarius*, *Ruminococcus torques*, *Bacteroides faecis,* and *Burkholderiales bacterium_1_1_47* were protective for DR. Consistent with our results, previous studies have also revealed a positive correlation of *Collinsella* with type 2 diabetes [36, 37]. *Collinsella* belongs to the family *Coriobacteriaceae*, which is usually considered as pathogens. They could affect host metabolism by stimulating gut leakage, modulating lipid metabolism and increasing cumulative inflammatory burden [38]. A recent study [39] has also confirmed that *Collinsella* abundance is positively correlated with circulating insulin in overweight and obese pregnant women with low dietary fiber intake, which may contribute to insulin resistance during pregnancy. In line with accumulating studies suggesting an involvement of *Collinsella* in the development of diabetes, our results first provided supportive evidence that there is a causal contribution of *Collinsella* to promote the development of DR.

Moreover, a growing number of studies have also reported that various other gut microbial taxa may contribute to DR risk. For example, a case-control study [40] has revealed that patients with DR and DM are characterized in enrichment of *Bifidobacterium* and *Lactobacillus* and depletion of *Escherichia-Shigella*, *Faecalibacterium*, *Eubacterium_hallii_group,* and *Clostridium*, compared to healthy individuals. Pasteurellaceae was found to be a biomarker with strong precision that distinguishes the DR patients from the DM group. Shang et al. [36] found that DR patients had higher abundance of *Oscillospira*, but lower abundance of *Megamonas* in a Chinese population. Given that dysbiosis of gut microbiome has been widely observed in patients with DR, reconstruction gut microbiome was thought to be able to prevent the development of DR. Recently, Beli et al. [14] reconstructed gut microbiome of mice by intermittent fasting and found that the altered gut microbiome (enrichment in *Firmicutes* and reduction in *Bacteroidetes*) produced more beneficial secondary BAs (e.g., TUDCA), which facilitate to decrease DR risk. The reduction of *Bifidobacterium*, *Clostridium*, and *Bacteroides* in *db/db*-IF mice was proposed to be responsible for the abnormal changes in levels of conjugated to unconjugated secondary BAs.

Several metabolic risk factors, including hyperglycemia, hypertension, and dyslipidemia, were known to confer higher risk for the progression of DR [10]. According to the Diabetes Control and Complications Trial (DCCT) [41], diabetes control with the goal of achieving blood glucose levels as close to the nondiabetic range as safely possible remarkably reduced the risk of the initiation and progression of DR, highlighting the importance of hyperglycemia in the etiology of DR. Hyperglycemia generally cause oxidative stress and glucose-mediated endothelium dysfunction and further influence retinal metabolic abnormalities [42]. For hypertension, epidemiological evidence has established that hypertension is a risk factor for retinopathy [43]. After 7 years’ follow up, there was a 47% reduction in risk of a decrease in vision with the 10 mm Hg reduction in SBP and 5 mm Hg reduction in DBP [44, 45]. In line with the concept that raised blood pressure (SBP, DBP) and blood glucose (2hGlu, FG, HbA1c) increased the risk of DR [10], our two-sample MR results provide robust genetic evidence to support that metabolic factors, such as hypertension, SBP, DBP, 2hGlu, FG, HbA1c, TG, and BMI, convey causal risk to DR.

In addition, our results also showed that BMI was causally associated with increased risk of DR based on European population. Consistently, previous studies based on cohorts of Caucasians [46] and Australians [47] have documented that there is a significant positive association between BMI and DR. However, other studies have reported a contradictory result that BMI is identified to be inversely associated with DR in cohorts of Singapore [48] and China [49]. This inconsistent finding indicates that the causal relationship between BMI and DR may be distinct in different ethnicities.

The associations of DR with serum lipids remained to be controversial. While HDL-C has been widely considered as a protective factor for DR [50–52], the NO BLIND study of Italy has reported that HDL-C is associated with a high risk of DR [53]. A similar contradictory association was also observed between total cholesterol and DR [54]. For TG and LDL-C, several studies have suggested that they are risk factors for DR, while others reported opposite results [54–56]. The Singapore Malay Eye Study reported higher total cholesterol levels as a protective factor for DR. In the current investigation, our large-scale two-sample MR results supports the protective effects of HDL-C and LDL-C on DR, and the detrimental role of TG on DR. IL-16 is a pro-inflammatory pleiotropic cytokine that functions as chemoattractant and modulator of T cell activation. It has been proposed that IL-16 in vitreous contributes to leukostasis and microvascular damage in the progression of proliferative diabetic retinopathy (PDR) [8] and rhegmatogenous retinal detachments (RRD) [57]. To date, no study on the association between serum IL-16 and DR has been reported. In the present study, we found a significant protective effect of serum IL-16 on DR by leveraging two-sample MR analyses, which is in contrast with previous findings in vitreous. This may be due to totally different mechanisms of serum and vitreous IL-16 on DR. The prototypical acute-phase protein CRP is a risk inflammatory biomarker for diabetes [58], and has been reported to be positively correlated with severity of DR [59]. However, inverse association of CRP with DR had also been documented [6]. According to two-sample MR analysis in the current study, we did not observe significant causal association of DR with CRP. In addition, although serum inflammatory cytokines TNF-α and IL-6 have been proposed to be biomarkers for DR by previous studies [9, 60], we did not find any significant evidence for causal association of DR with them in our present investigation.

Finally, in the mediation analysis, *Streptococcus salivarius* was discovered to be causal gut microbe for DR through mediating FG. *S. salivarius,* which is a member of *viridans streptococci*. It is one of the primary inhabitants of human intestine and oral, and usually plays benefical role but occasionally causes opportunistic infection. In the oral cavity, colocalized *S. salivarius* inhibits the emergence of pathogens by producing bacteriocins, such as lantibiotics. Thus, *S. salivarius* K12 is now used as an oral probiotic worldwide to sustain the hemostasis of oral microbiome [61]. The immunomodulatory and anti-inflammatory effect of *S. salivarius* was reported in intestinal epithelial cells [62] and mouse models [63]. Although the association of *S. salivarius* with DR has not been reported yet, the anti-diabetic effect of *S. salivarius* has been documented [64]. Chen et al. [64] found that γ-Aminobutyric acid (GABA) produced by *S. salivarius* subspecies *thermophiles* fmb5 in yogurt fermentation was capable of improving hyperglycaemia and decreasing the concentrations of serum total cholesterol and triacylglycerol. This supports our conclusion that intestinal *S. salivarius* negatively regulate blood glucose to decrease the risk of DR. In addition, in view of multiple lines of evidence [65–73] have reported that combining multi-omic data, including GWAS, single-cell RNA sequencing, and epigenetic data, contribute to reveal the molecular mechanism of complex diseases. More integrative genomic analyses of microbiome with other omics are needed to disentangle the casual etiology of DR.

In conclusion, our study proposed a causal association between *S. salivarius* and DR. Genetically elevated level of *S. salivarius* is causal associated with decreased risk of developing DR. Moreover, the causal effect of *S. salivarius* partially mediated by reducing level of fasting glucose. *S. salivarius* seems to be a possible probiotic supplement that could be used in the treatment or prevention of DR. Therefore, our findings support the hypothesis that gut microbes could be causal factors for DR and highlight the need for extensive research on how gut microbes affect the etiology of DR.

## Data Availability

All the GWAS summary statistics used in this study can be accessed in the official websites (https://gwas.mrcieu.ac.uk/). The GTEx eQTL data (version 8) were downloaded from Zenodo repository (https://zenodo.org/record/3518299#.Xv6Z6igzbgl). The code to reproduce the results is available in a dedicated GitHub repository (https://github.com/SulabMR/207GM_DR_MR).

https://zenodo.org/record/3518299#.Xv6Z6igzbgl

## Declarations

## Authors’ contributions

Y.M., J.L., and J.S. conceived and designed the study. J.L., G.Z., C.D., Y.Z., and Y.M. contributed to management of data collection. J.L., Y.M., and G.Z., conducted bioinformatics analysis and data interpretation. Y.M., J.S., and J.L. wrote the manuscripts. All authors reviewed and approved the final manuscript.

## Ethics approval and consent to participate

Not applicable

## Consent for publication

Not applicable

