## Supplementary figures and images for "Mendelian randomization analysis identifies a causal effect of Streptococcus salivarius on DR mediating via the level of host fasting glucose"

### Supplementary Figure S1

Supplementary Fig. 1

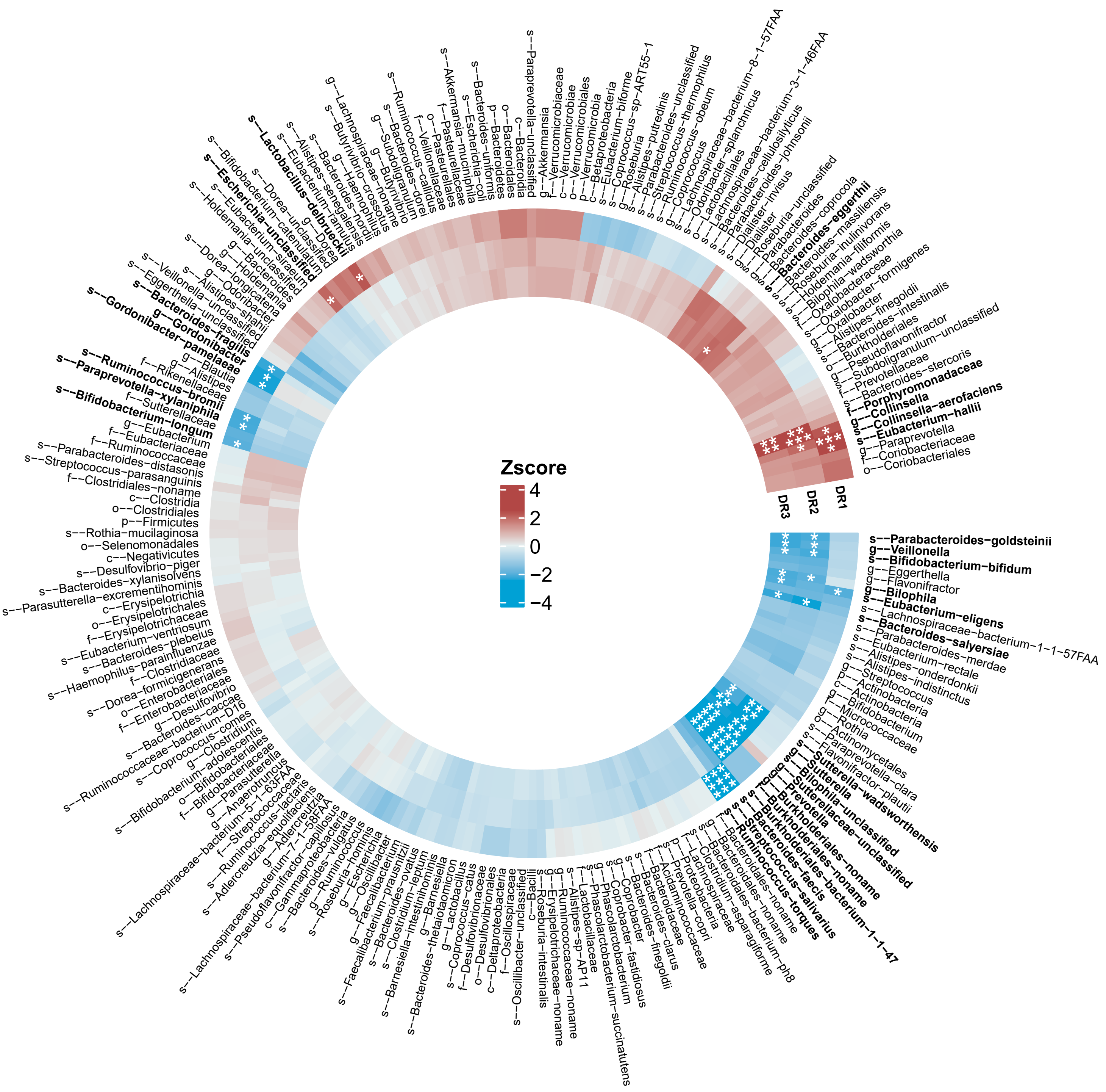

### Supplementary Figure S2

#### Supplementary Fig. 2

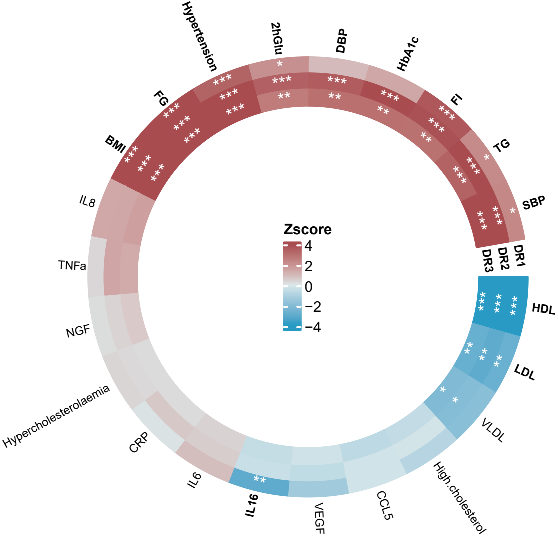
